# Systematic Evaluation of Adiposity Indices as Predictors of Hypertension Risk in Children and Adolescents

**DOI:** 10.1101/2024.11.26.24318031

**Authors:** Bowen Zhu, Shumin Zhan, Hui Shi, Xingyun Wang, Jingwen Yue, Jianfang Gao, Tongshuai Wang, Rui Wang, Xirong Guo, Junfen Fu

## Abstract

**Objective:** To systemically examine the concomitant impact of adiposity indices on hypertension in children and adolescents.

**Methods:** A community-based cohort study by China Health and Nutrition Survey (2009-2011) included 262 children and adolescents aged 7-17. Anthropometric and lipid profiles were analyzed for hypertension risk using logistic and cross-lagged models.

**Results:** Of the 227 participants (mean age, 16.5 [2.3] years), 147 (53.9%) were boys. Over a two-year period, 26.4% developed hypertension, 5.3% had elevated systolic blood pressure (SBP), and 25.6% had elevated diastolic blood pressure (DBP). After adjusting for covariates, higher body mass index (BMI), waist circumference (WC), hip circumference (HC), triceps skinfold thickness (TST), and body roundness index (BRI) were associated with increased hypertension risk. Incorporating these measures into a BP-based model improved the AUC for hypertension prediction from 0.551 (95% CI: 0.463, 0.640) to 0.670 (95% CI: 0.592, 0.748). A longitudinal relationship were observed between these adiposity indices and hypertension symptoms.

**Conclusions:** The study suggests anthropometry parameters are superior to blood lipid indicators in predicting the occurrence of hypertension in children. Abdominal obesity, as indicated by WC, HC, and BRI, accounts for a significant portion of the risk for hypertension, particularly in children aged 7 to 12 years.

## 1. Introduction

Hypertension affects about 4% of children and 11% of adolescents globally [1]. The early identification and proper intervention of high blood pressure or hypertension during childhood are extremely crucial because these conditions are related to cardiovascular morbidity (left ventricular hypertrophy and high carotid intima-media thickness) and mortality in adulthood and are commonly associated with obesity [2–4]. A comprehensive analysis of 2033 studies spanning 154 countries has revealed a significant prevalence of overweight and obesity among children and adolescents[5]. The pooled estimates indicate that 8.5% are overweight and 14.8% are obese, which translates to roughly one in every five young individuals facing the challenge of excess weight. China’s rapid economic progress has brought about major lifestyle and societal transformations, resulting in a surge in metabolic disorders. This trend is likely to have a substantial effect on the cardiovascular disease (CVD) burden in the country. The number of children aged 5 to 19 years with obesity is predicted to rise to 254 million by 2030 [6]. It is believed that obesity-related primary hypertension has contributed to the rise in hypertension prevalence, which has doubled from 1% to 4% over the past three decades[7–8].

Enhancing our comprehension of the anthropometric and biochemical indicators of adiposity profiles in the context of hypertension is essential for the prevention and management of this condition in children and adolescents. In a cohort study of youths, a high normal body weight above the 60th percentile of BMI for age was associated with an increased risk of hypertension [9]. The suggested thresholds of 0.50 and 0.46 for two distinct pediatric groups have demonstrated robust predictive capabilities for identifying individuals with two or more cardiometabolic risk factors in populations from six countries [10]. Furthermore, innovative anthropometric indices such as the Body Shape Index (BSI), Body Roundness Index (BRI), Conicity Index (CI), and Visceral Fat Index (VFI) have been introduced to more accurately pinpoint subcutaneous and visceral fat distribution. A meta-analysis with 21108 children and adolescents confirmed that original ABSI and modified ABSI do not perform as well as traditional anthropometric indicators, such as BMI and WC. BMI continues to be the most effective indicator for screening high blood pressure (HBP) in pediatric populations[11]. Rui Chen et al. reported that WHtR and BRI can be recommended to identify hypertension, dyslipidemia, abdominal obesity, and clustered cardio-metabolic risk factors (CMRFs) in 7-17-year-old teenagers [12]. However, the majority of studies have been conducted with adult populations, and the interplay and individual impact of biochemical measures, anthropometric data, and composite metrics on hypertension remains to be fully elucidated. Furthermore, there is a scarcity of data that can differentiate between the hypertension risk in childhood associated with initial obesity-related metrics and the predictive capacity. In this study, we aimed to systemically investigate the temporal and bidirectional relationship between anthropometric and biochemistry measures of obesity and hypertension symptoms and further explore the prediction ability of obesity-related measures for hypertension status using a large cohort.

## 2. Materials and Methods

### 2.1 Study cohorts

This survey encompassed children and adolescents aged 7 to 17 years, with two follow-up visits conducted between 2009 and 2011. Valid blood pressure (BP) data, comprising three repeated readings, were obtained. Participants with missing BP values during the follow-up period were excluded from the analysis. The data was sourced from the China Health and Nutrition Survey (CHNS), an ongoing, open cohort study. The CHNS is a comprehensive, nationwide, prospective cohort study that includes a representative sample of the Chinese population. Its primary objective is to assess the impact of health, nutrition, and family planning policies and programs implemented by both national and local governments. Further details regarding the study design and data collection methodologies of the CHNS have been documented in prior publications [13]. Blood samples were collected in the 2009 year, and data information on biochemical indicators was obtained. Since blood samples were collected in 2009 year, a total of 768 participants with blood biomarkers information were collected in 2009. After excluding those who had 3-day high protein intake (n=17), elevated SBP or DBP (defined as age- and sex-specific 90th percentile, n=221), glucose above 7.0 mmol/L or HbA1c above 6.5% (n=8), BMI above 35 kg/m^2^ (n=1), missing data of systolic BP or diastolic BP (n=133), 388 participants were included in the formal analysis. After that, a total of 161 participants were excluded as they had lost visits during the follow-up duration (***Supplementary Figure 1***). Demographic and behavioral characteristics of participants included in the final analysis (n=227) were compared with those of excluded participants (***Supplementary Table 1***). All data and samples were collected after written informed consent was obtained from parents or caregivers for children and adolescents aged 7 to 17 years. The CHNS was granted approval by the Institutional Review Board at the University of North Carolina at Chapel Hill, as well as by the local Institutional Review Board of Ethics Committee.

### 2.2 Data collection

A standardized structured questionnaire was administered by trained health staff to collect socio-demographic variables (in 2009) including age, gender, urban-rural residence, soft/sugared fruit drinks, total protein intake, total fat intake, total carbohydrate intake, and total energy intake. Physical examinations of waist circumference (WC), hip circumference (HC), height, weight, triceps skinfold thickness (TST), upper arm circumference (UAC), and BP were performed by trained clinical staff. All individuals maintained a regular life pattern for at least three days before blood sample collection and 12 ml of blood was collected (in three 4 ml tubes) on an empty stomach. Biomarker data collected in CHNS 2009 involves the release of 26 fasting blood measures on individuals aged 7 and older [14]. The estimated glomerular filtration rate (eGFR) was calculated using the Schwartz formula [15]. The individuals were categorized into groups, namely, residence, urban, and rural. Individual dietary intake for 3 consecutive days was determined for every household member. This step has been achieved by asking individuals each day to report all food consumed away from home on a 24-hour recall basis, and the same daily interview has been used to collect at-home individual consumption. Soft/sugared fruit drinks information was assessed by the question including “Drink soft/sugared fruit drinks?” or “How often drink soft/sugared fruit drinks?”.

### 2.3 Adiposity Indices Measures

Adiposity indices measures included body mass index (BMI), waist circumference (WC), hip circumference (HC), triceps skinfold thickness (TST), waist to hip circumference ratio (WHR), upper arm circumference (UAC), triglyceride (TG), high-density lipoprotein cholesterol (HDL-c), blood glucose, serum uric acid (SUA), conicity index (CI), body shape index (BSI), visceral fat index (VFI), body roundness index (BRI). BMI was calculated as weight(kg)/height^2^(m^2^) and was converted into age and gender-specific BMI percentiles [16]. Abdominal obesity was defined as waist circumference age- and sex-specific 90th percentile, determined by the cut-off points of standards for Chinese children and adolescents [17]. Waist to height ratio (WHR) was calculated as waist circumference(cm)/ height(cm). The calculation of the novel anthropometric indices is described below, with the relevant reference cited at the end of each formula:

CI = WC/0.109* (Wt/Ht)^1/2^ [18]
BSI = WC∗Wt^−2/3^∗Ht^-5/6^ [19]
VFI=WC/ [39. 68 + 1.88*BMI*TG/1. 03] * [1.31/HDL-C] (for male),
VFI=WC/(36. 58+1. 89*BMI)*(TG/0. 81)*(1. 52/HDL-C) (for female) [20]
BRI = 364.2−365.5∗ (1 − ((0.5∗WC/π)^2^/(0.5∗Ht)^2^))^0.5^ [21]
The unit of TG, HDL-c, and LDL-c was mmol/L.

TG was measured using the GPO-PAP (Hitachi 7600, Kyowa, Japan). HDL-c was measured using the Enzymatic method (Hitachi 7600, Kyowa, UK). Glucose was measured using the GOD-PAP (Hitachi 7600, Randox, UK).

### 2.4 Diagnostic Criteria and Definitions of Outcome

Systolic and diastolic blood pressure values were determined based on the average of three separate measurements. Participants’ blood pressure status, classified as either normal or elevated, was ascertained using sex- and age-specific cutoff values that are applicable to Chinese school-age children and adolescents. In accordance with Cook’s criteria, elevated blood pressure is defined as a systolic or diastolic blood pressure equal to or greater than the 90th percentile for age, sex, and height [22]. The evaluation of blood pressure outcomes was conducted using three distinct categorization methods: hypertension (defined by elevated SBP or elevated DBP) versus non-hypertension; elevated SBP versus normal SBP; and elevated DBP versus normal DBP.

### 2.5 Statistical Analysis

Data were presented as mean ± standard deviation (SD) and median with interquartile range (IQR) for continuous variables or number (percentage), for categorical variables. Data on demographics, anthropometric and biochemistry measures were compared using the chi-square test, and Fisher’s exact test for categorical variables. The Wilcoxon rank-sum test was performed for continuous variables. Multivariable logistic regression models were employed to evaluate the longitudinal associations of adiposity indices with incident hypertension, elevated SBP, and elevated DBP, respectively. Multivariable models were sequentially adjusted for (1) SBP, and/or DBP; (2) plus age, and gender; and (3) plus baseline residence, eGFR, soft fruit drinks, and total carbohydrate intake. The areas under the receiver operator characteristics curve (AUC) were calculated to assess the prediction ability of adiposity indices for hypertension, elevated SBP, and elevated DBP. Continuous net reclassification improvement (NRI), and absolute integrated discrimination improvement (IDI) were used to assess whether adding the obesity-related measures could improve risk discrimination and reclassification for hypertension, elevated SBP, and elevated DBP prediction above the baseline SBP, /or DBP. The cross-lagged panel models were applied to analyze the temporal relationship between obesity-related measures (as continuous variables) and SBP, and/or DBP levels (as categorical variables). The lagged effect between the observed variables, the correlation between the selected adiposity indices (BMI, CI, WC, HC, TST and UAC, BRI) at each phase, and the residual variance of the observed variables (SBP, and/or DBP levels) were estimated in the cross-lagged panel analysis, which elucidates the temporal relationship between correlated variables and indicates the precursor variable. Several sensitivity analyses were performed to test the robustness of our findings. First, we treated obesity-related measures, SBP, and DBP as continuous variables, to determine the linear associations between adiposity indices and BP. Prespecified subgroup analyses were conducted based on age (7-12 years and 13-17 years), and gender (boys and girls) in secondary analyses. *P* < 0.05 (two-sided) was considered statistically significant. All analyses were conducted with SAS software (v.9.4, SAS Institute Inc., Cary, NC, USA).

## 3. Result

### 3.1 Baseline Characteristics of Pediatric and Adolescent Participants Across Various Systolic and Diastolic Blood Pressure Groups

Among the 227 participants included in the present analysis (64.8% of whom were boys), the mean age was 11.7 (SD, ±2.3) years. During a median of 2 years of follow-up, 60 cases of incident hypertension (26.4%), 12 cases of incident elevated SBP (5.3%), and 58 cases of elevated DBP (25.6%) were identified. Children diagnosed with hypertension exhibited significantly higher measurements for WC, HC, BMI, TST, and BRI. Additionally, a higher proportion of these children were girls and were classified as overweight. Compared with the normal SBP group, the children and adolescents diagnosed with elevated SBP had a higher level of waist circumference, hip circumferences, BMI, TST, BRI, ferritin, total protein, as well as more proportions of overweight. Compared with the normal DBP group, the children and adolescents diagnosed with elevated DBP had a higher level of WC, HC, BMI, TST, low level of total carbohydrate intake, as well as more proportions of girls, overweight (***Table 1***).

**Table 1.**
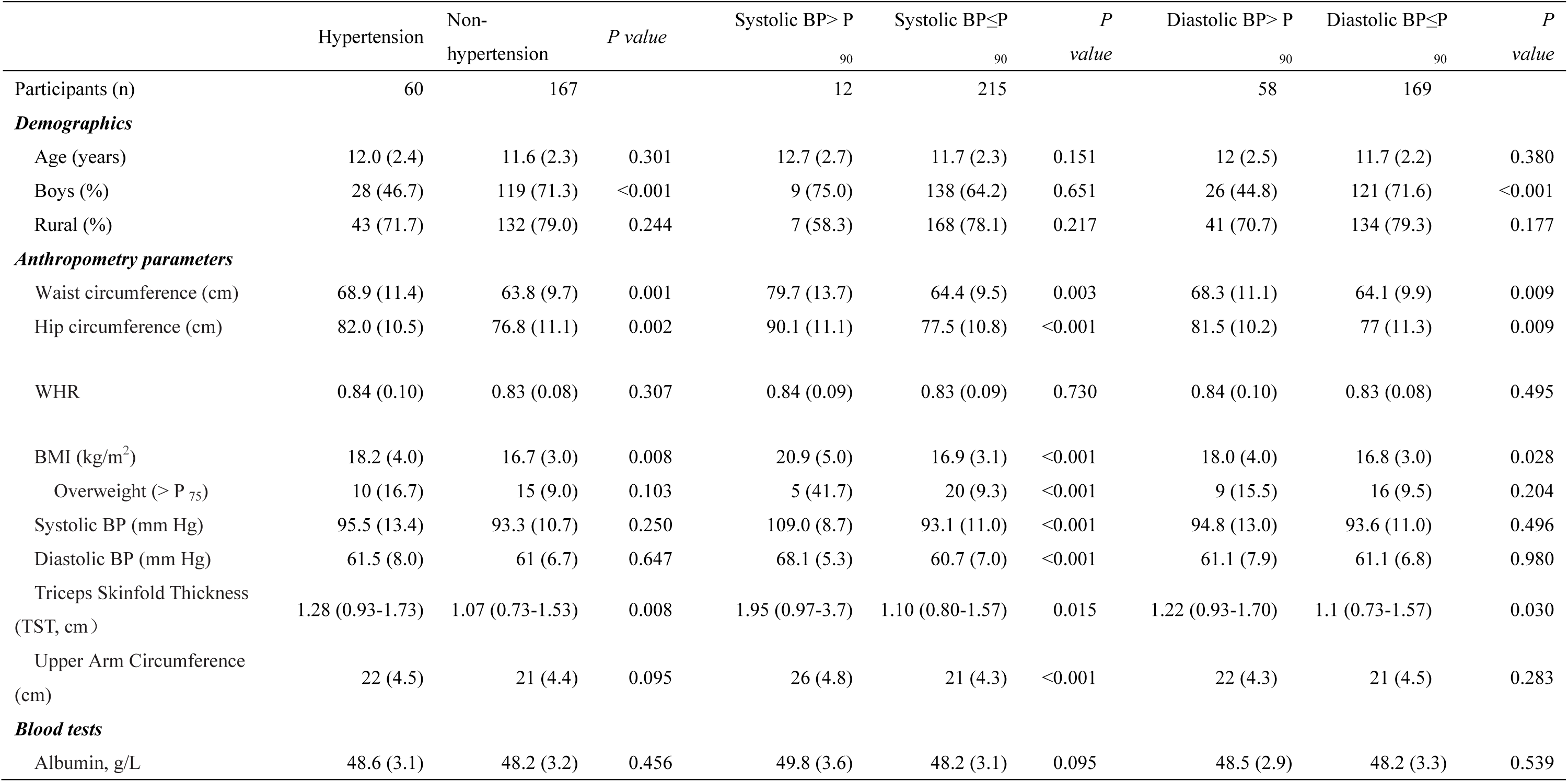
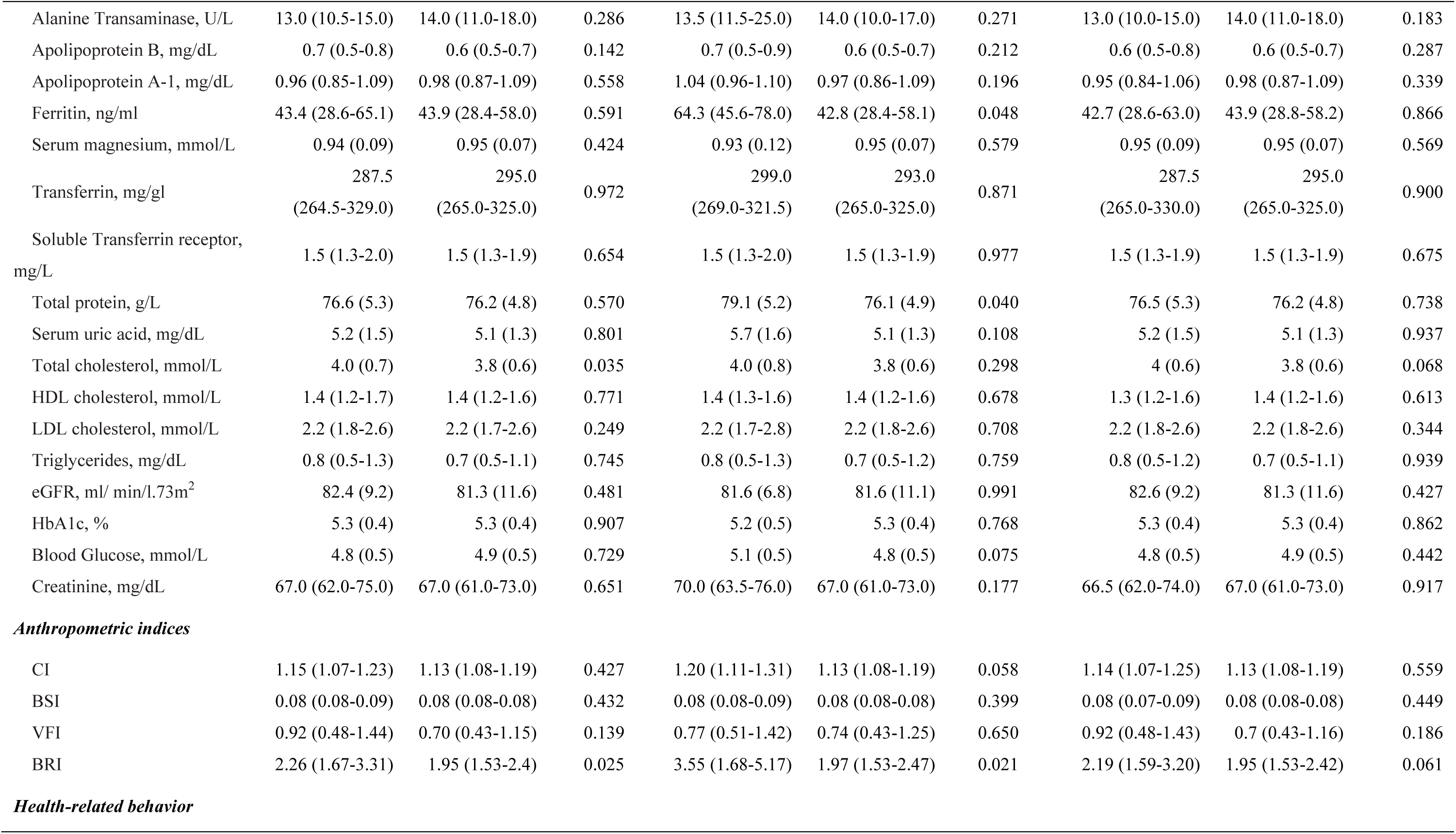
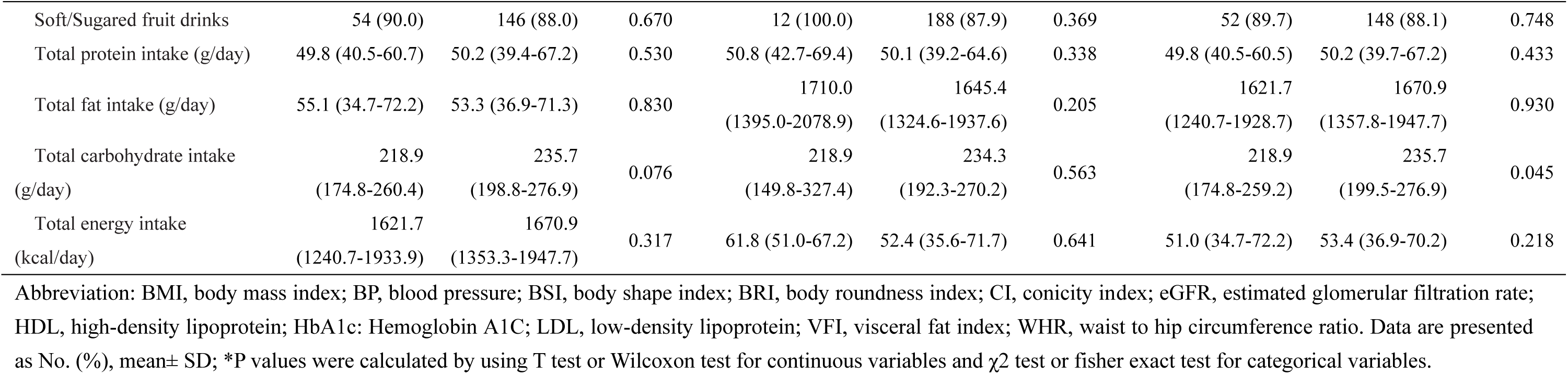
Baseline characteristics of the individuals stratified by Hypertension, Systolic BP and Diastolic BP in the CHNS dataset (n=262)

### 3.2 Correlations Between Body Adiposity Measures and the Onset of Hypertension

The risk ratios (RRs) and 95% confidence intervals (CIs) of incident hypertension for each adiposity index (per SD) are shown in Figure 1. After adjusting for covariates, BMI, WC, HC, TST, and BRI were associated with a higher risk of hypertension. When defining the outcome as elevated SBP (SBP > P_90_), BMI, WC, HC, TST, UAC, SUA, CI, and BRI were associated with a higher risk of elevated SBP. While, BMI, WC, HC, TST, TG, and BRI were associated with a higher risk of elevated DBP (***Figure 1***).

**Figure 1.**
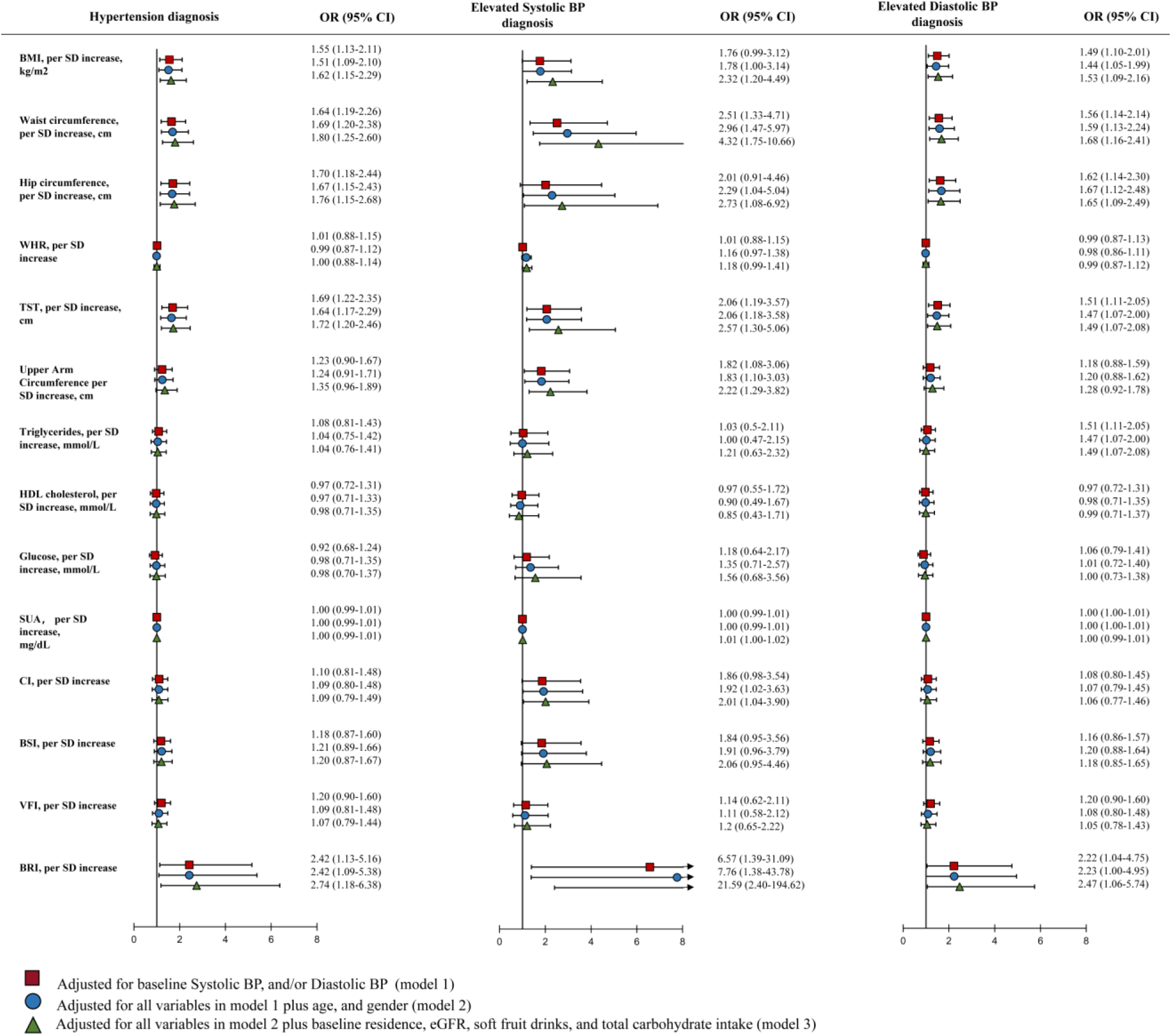
Associations between anthropometric indices/metabolic measures and hypertension diagnosis (Abbreviations: BMI, body mass index; BRI, body roundness index; CI, conicity index; TST, triceps skinfold thickness; WHR, waist-to-hip ratio. * 0.01<P≤0.05; ** 0.001<P≤0.01; *** P<0.001)

### 3.3 The Incremental Predictive Value of Adiposity Indices in Assessing the Risk of Hypertension, High Systolic Blood Pressure, and High Diastolic Blood Pressure

A total of seven adiposity indices were identified, including BMI, WC, HC, TST, UAC, BRI, and CI. The inclusion of HC and BRI was found to enhance the net reclassification ability and the overall predicted probability for hypertension. Similarly, incorporating WC was shown to improve the net reclassification ability and the overall predicted probability for elevated SBP. Adding the WC, HC, and BRI could improve the net reclassification ability and overall predicted probability for elevated DBP. Furthermore, by incorporating BMI, WC, and HC into the basic model—which considers continuous SBP and DBP — the area under the curve (AUC) for hypertension prediction was significantly enhanced, increasing from 0.551 (95% CI: 0.463, 0.640) to 0.670 (95% CI: 0.592, 0.748). For SBP prediction, the AUC remained high at 0.873, with a CI ranging from 0.771 to 0.974. Similarly, for elevated DBP prediction, the AUC improved from 0.531 (95% CI: 0.440, 0.621) to 0.649 (95% CI: 0.570, 0.729) (***Table 2)***.

**Table 2.** Performance of Risk Prediction Models for 2-Year Risk of Incident Hypertension by Including selected Anthropometry parameters and metabolic Measures.

|  | AUC (95% CI) | Difference (95% CI) | NRI (95% CI) | Absolute IDI (95% CI) |
| --- | --- | --- | --- | --- |
| **2-Year diagnosis of Hypertension** |  |  |  |  |
| Basic model<sup>a</sup> | 0.551 (0.463, 0.640) | Reference | Reference | Reference |
| Basic model+BMI | 0.624 (0.542, 0.707) | 0.073 (-0.014, 0.161) | 0.165 (-0.120, 0.451) | 0.036 (0.005, 0.066)* |
| Basic model+Waist circumference | 0.641 (0.559, 0.724) | 0.090 (-0.012, 0.192) | 0.244 (-0.044, 0.531) | 0.044 (0.013, 0.075)** |
| Basic model+Hip circumference | 0.646 (0.563, 0.729) | 0.095 (-0.001, 0.190) | 0.468 (0.189, 0.748)* | 0.040 (0.013, 0.066)** |
| Basic model+TST | 0.625 (0.542, 0.707) | 0.073 (-0.029, 0.176) | 0.148 (0.076, 0.658)* | 0.027 (-0.009, 0.063) |
| Basic model+BRI | 0.605 (0.518, 0.692) | 0.054 (-0.048, 0.156) | 0.360 (0.069, 0.651)* | 0.041 (0.007, 0.074)* |
| Basic model+BRI, Hip circumference | 0.655 (0.574, 0.735) | 0.103 (0.003, 0.204)* | 0.527 (0.241, 0.812)* | 0.056 (0.020, 0.092)** |
| Basic model+BMI, Waist circumference, Hip circumference | 0.670 (0.592, 0.748) | 0.119 (0.021, 0.217)* | 0.455 (0.168, 0.742)** | 0.051 (0.018, 0.084)** |
| **2-Year diagnosis of Elevated Systolic BP** |  |  |  |  |
| Basic model<sup>b</sup> | 0.873 (0.771, 0.974) | Reference | Reference | Reference |
| Basic model+BMI | 0.889 (0.795, 0.982) | 0.016 (-0.026, 0.058) | 0.243 (-0.324, 0.818) | 0.051 (0.034, 0.137) |
| Basic model+Waist circumference | 0.896 (0.797, 0.995) | 0.023 (-0.036, 0.082) | 0.700 (0.153, 1.249)* | 0.113 (0.001, 0.225)* |
| Basic model+Hip circumference | 0.885 (0.783, 0.987) | 0.013 (-0.029, 0.054) | 0.209 (-0.365, 0.782) | 0.037 (-0.02, 0.094) |
| Basic model+TST | 0.885 (0.779, 0.991) | 0.013 (-0.048, 0.073) | 0.413 (-0.160, 0.986) | 0.144 (-0.031, 0.318) |
| Basic model+Upper Arm circumference | 0.888 (0.801, 0.975) | 0.015 (-0.026, 0.057) | 0.784 (0.277, 1.290)* | 0.048 (-0.018, 0.114) |
| Basic model+CI | 0.888 (0.777, 0.999) | 0.017 (-0.030, 0.063) | 0.340 (-0.240, 0.919) | 0.038 (-0.030, 0.105) |
| Basic model+BRI | 0.886 (0.766, 1.000) | 0.014 (-0.049, 0.077) | 0.711 (0.142, 1.280)* | 0.158 (-0.011, 0.328) |
| Basic model+BRI, Hip circumference | 0.893 (0.776, 1.000) | 0.021 (-0.039, 0.081) | 0.636 (0.066, 1.207)* | 0.157 (-0.01, 0.325) |
| Basic model+BMI, Waist circumference, Hip circumference | 0.897 (0.797, 0.996) | 0.024 (-0.034, 0.083) | 0.710 (0.163, 1.258)* | 0.113 (0.001, 0.224)* |
| **2-Year diagnosis of Elevated Diastolic BP** |  |  |  |  |
| Basic model<sup>c</sup> | 0.531 (0.440, 0.621) | Reference | Reference | Reference |
| Basic model+BMI | 0.605 (0.523, 0.687) | 0.108 (-0.027, 0.242) | 0.200 (-0.089, 0.490) | 0.044 (0.015, 0.072)** |
| Basic model+Waist circumference | 0.623 (0.537, 0.709) | 0.125 (-0.012, 0.263) | 0.333 (0.039, 0.628)* | 0.037 (0.009, 0.065)** |
| Basic model+Hip circumference | 0.631 (0.547, 0.714) | 0.134 (0.008, 0.259)* | 0.456 (0.175, 0.738)** | 0.043 (0.014, 0.072)** |
| Basic model+TST | 0.597 (0.511-0.683) | 0.100 (-0.042-0.242) | 0.322 (0.027,0.618)* | 0.013 (-0.021-0.046) |
| Basic model+BRI | 0.575 (0.480, 0.670) | 0.078 (-0.072, 0.228) | 0.349 (0.054,0.643)* | 0.052 (0.017, 0.087)** |
| Basic model+BRI, Hip circumference | 0.639 (0.556, 0.721) | 0.141 (0.006, 0.277)* | 0.577 (0.292, 0.861)** | 0.037 (0.009, 0.065)** |
| Basic model+BMI, Waist circumference, Hip circumference | 0.649 (0.570, 0.729) | 0.152 (0.022, 0.282)* | 0.349 (0.054, 0.643)** | 0.044 (0.014, 0.073)** |

Abbreviations: AUC, area under curve; BMI, body mass index; BRI, body roundness index; CI, conicity index; eGFR, estimated glomerular filtration rate; TST, triceps skinfold thickness. a Basic model: baseline age, gender, systolic BP, diastolic BP, and eGFR. b Basic model: baseline age, gender, systolic BP, and eGFR; c Basic model: baseline age, gender, diastolic BP. \* 0.01<P≤0.05; \*\* 0.001<P≤0.01; \*\*\* P≤0.001

### 3.4 The Temporal Dynamics of the Association Between Body Adiposity Measures and Elevated Systolic/Diastolic Blood Pressure

The fully cross-lagged model was used when compared to both the adiposity indices measures to BP levels (defined as elevated SBP or elevated DBP) and the BP levels to the obesity-related measures (***Figure 2***). Of note, there existed no directional associations between the most obesity-related measures and any hypertension symptoms. Baseline adiposity indices including WC, HC, TST, UAC, BRI, and CI were unidirectional longitudinally associated with hypertension symptoms (elevated SBP or elevated DBP). Cross-lagged path estimates showed that only BMI was directional and longitudinally associated with hypertension symptoms. Baseline SBP (in the 2009 year) was longitudinally negatively associated with later BMI (in 2011 year), while baseline DBP (in 2009 year) was significantly positively associated with BMI (in 2011 year).

**Figure 2.**
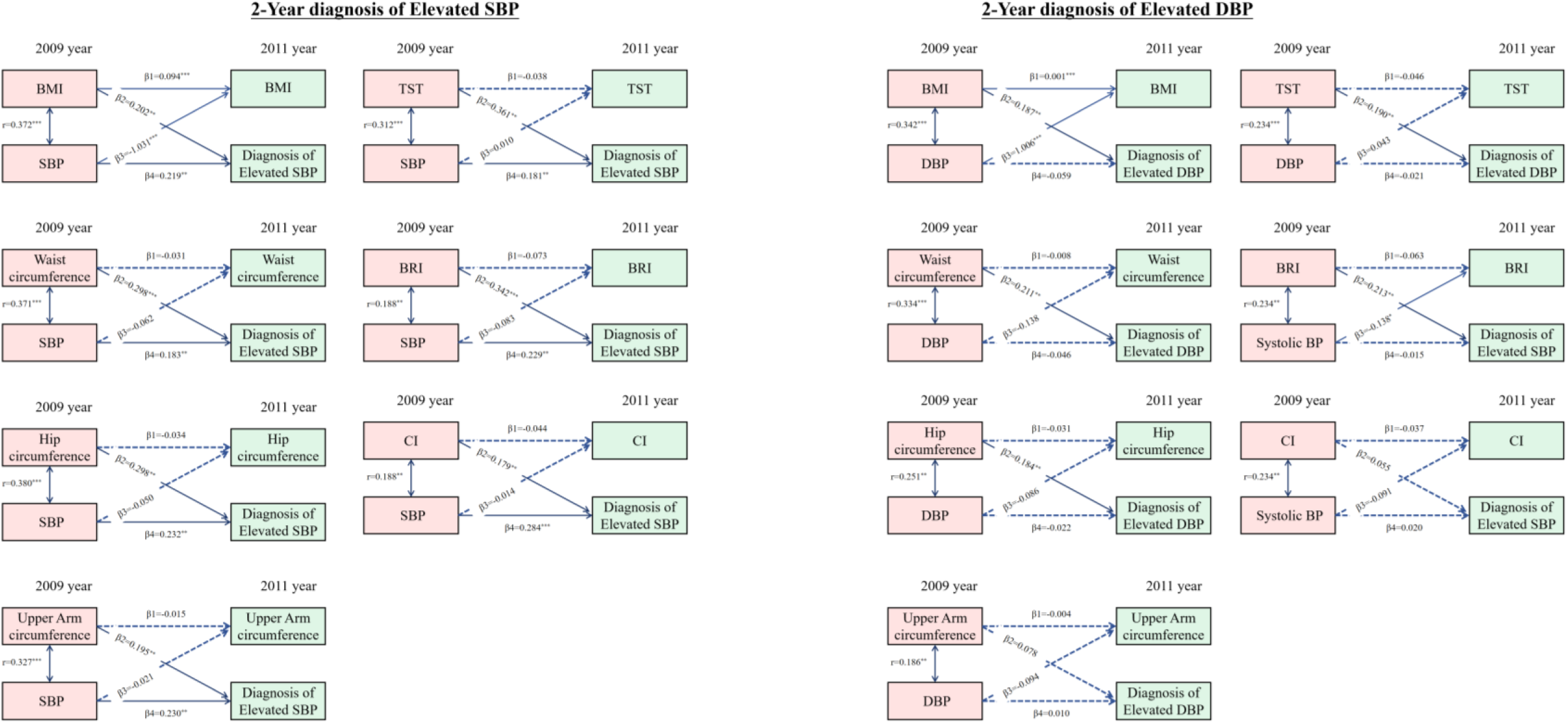
Estimates from the cross-lagged model for the associations between anthropometry parameters and incident hypertension classified by elevated SBP and elevated DBP (Abbreviations: BMI, body mass index; BRI, body roundness index; CI, conicity index; eGFR, estimated glomerular filtration rate; TST, triceps skinfold thickness. All estimates were standardized. * 0.01<P≤0.05; ** 0.001<P≤0.01; *** P<0.001).

### 3.5 Sensitivity and Subgroup Analysis

In the sensitivity analysis, the linear analysis revealed significant longitudinal associations between various adiposity indices including BMI, WC, HC, TST, UAC, BSI, and BRI, with a higher risk of SBP and DBP, treated as continuous variables **(*Supplementary Figure 2*)**. In subgroup analysis, the significant associations of BMI, WC, HC, TST, and BRI with risk of hypertension only existed in the younger age group (7∼12 years) after accounting for multiple variables. In the subgroup classified by gender, the significant associations of BMI with the risk of hypertension only existed in girls. Whereas, the associations of UAC, and high level of TG with risk of hypertension only existed in boys **(*Supplementary Table 2 and Supplementary Table 3*)**.

## 4. Discussion

In this cohort study of the resident Chinese pediatric and adolescent population, we identified a significant correlation between elevated BMI, WC, HC, TST, and BRI with the prevalence of hypertension. This association remained consistent, regardless of initial systolic and diastolic blood pressure measurements, demographic variables, lifestyle factors, and eGFR. In contrast, we observed no significant link between hypertension and UAC, triglycerides (TG), HDL-c, blood glucose levels, serum uric acid (SUA), CI, BSI, and VFI. The relationships of these adiposity indices with both elevated SBP and DBP mirrored their associations with hypertension, with the exception that an increased UAC and CI were linked to a higher probability of elevated SBP. It is noteworthy that among the five adiposity indices, only BMI demonstrated a consistent, directional longitudinal association with the symptoms of hypertension.

Previous research has consistently highlighted the relationship between adiposity indices and pediatric hypertension [23,24]. Our study adds to the existing literature by providing evidence that an increased WC is positively correlated with elevated blood pressure in pediatric populations. The accumulation of abdominal fat is known to secrete various adipokines and cytokines that can lead to endothelial dysfunction and increased peripheral vascular resistance, thereby contributing to the development of hypertension [25–27]. Moreover, our study reveals that an expanded HC is associated with a higher degree of fat deposition in the buttocks and thighs, consequently increasing the likelihood of hypertension in the pediatric population. TST could serve as a valuable anthropometric marker for assessing cardiovascular risk in children. This association may be attributed to the secretion of adipose-derived hormones and inflammatory cytokines that can lead to endothelial dysfunction and heightened vascular resistance [28]. The areas under the ROC curves for WC and HC in predicting pediatric hypertension were higher than those for BMI. These findings suggested that body fat distribution, rather than overall obesity, maybe a more sensitive predictor of cardiovascular health in the pediatric population [29].

The areas under the receiver operating characteristic (ROC) curves for WC and HC in the prediction of pediatric hypertension were notably higher compared to those for BMI. These results suggest that the distribution of body fat, rather than overall adiposity, may be a more sensitive predictor of cardiovascular health in the pediatric population. BRI, which quantifies visceral fat and total body fat percentages, was significantly associated with elevated blood pressure in children [30].

In prior research, the BRI has been identified as a more effective predictor than other anthropometric measures for assessing the risk of a range of clinical outcomes in adults. These include conditions such as cardiometabolic disorders, renal impairment, and malignancies [31–35]. There is an emerging consensus that visceral fat poses a significantly greater health risk compared to subcutaneous fat, as it is more strongly associated with an increased risk of various diseases in adults [36–37]. While the concept that BRI can estimate total and regional fat percentages seems plausible and potentially superior in adults, the evidence linking BRI to diseases in children is currently limited. In the study, the area under the ROC curves for BRI in predicting pediatric hypertension was found to be lower compared to WC and HC. This suggests that BRI’s predictive power for pediatric hypertension may not be as effective as it is in adults.

Theoretically, assuming the shape of the body as an ellipse with the long axis height and the short axis WC, BRI can be calculated as the eccentricity of this ellipse via human modeling. Acknowledging the inherent disparities in body contours between the pediatric and adult demographics, it is crucial to appreciate that indices like the BRI may not be directly comparable across these groups. The efficacy of BRI as a predictor for hypertension could be subject to variation owing to the unique growth trajectories, body composition differences, and the influence of developmental milestones that are characteristic of the pediatric population. Therefore, the application of BRI in pediatric assessments should be approached with caution, and further research is needed to establish its validity and reliability in predicting hypertension specifically within the pediatric demographic. For practical reasons, there remains a need for a simple and effective children-specific indicator to better reflect visceral obesity. Our study revealed that UAC and CI were significantly correlated with systolic blood pressure but not with diastolic blood pressure. This distinction could be due to the different physiological mechanisms underlying the two types of blood pressure. Systolic blood pressure is more influenced by arterial stiffness, cardiac output, and abnormal changes in brain cortical structure, whereas diastolic blood pressure is more affected by peripheral vascular resistance [38,39]. The fact that our measurements are more strongly associated with SBP may indicate a closer link with factors affecting arterial stiffness, and brain cortical structure.

Our study’s findings suggest that in the pediatric population, there is no significant association between blood lipid levels (triglycerides, LDL-c, HDL-c, total cholesterol, SUA) and the development of hypertension. This observation challenges the widely held belief that dyslipidemia is a consistent predictor of cardiovascular risk, including hypertension in adults. The absence of a correlation in our pediatric cohort could be attributed to several factors. First, the pathophysiology of hypertension in children may differ fundamentally from that in adults, where lipid metabolism plays a more established role [40]. Children are in a dynamic phase of growth and development, with a rapidly changing metabolic profile that might not yet be fully aligned with the traditional risk factors seen in adulthood [41].

Additionally, the dietary and lifestyle factors that significantly influence lipid profiles in adults may not have had sufficient time to exert their effects in children. As children’s dietary habits and physical activity levels evolve, their lipid profiles may not have stabilized to a point where they can significantly influence blood pressure. The association between adiposity indices and hypertension in children appears to be age-dependent, with our study indicating a significant correlation within the 7 to 12 years age group, which was not observed in the 13 to 18 years age group. This age-specific association may reflect the dynamic changes in growth, development, and metabolic patterns that occur during childhood and adolescence. The physiological changes during adolescence, including growth spurts, hormonal fluctuations, and the maturation of various organ systems, could influence the relationship between adiposity indices and blood pressure in complex ways that are not yet fully understood. The distribution of body fat and its metabolic impact may be different in adolescents compared to adults, potentially mitigating the effects of excess adiposity on blood pressure regulation.

### Strengths and Limitations

A significant advantage of our study is its community-based approach, which successfully enrolled a substantial sample of children and adolescents aged 7 to 17 in China. This age group has been notably underrepresented in research pertaining to pediatric hypertension. Limitations include the possibility of residual confounding inherent to the observational design, including the possibility of differential distribution of unmeasured or incompletely measured confounders such as birth weight and gestational age. We can also not exclude limitations in the generalizability of the results in this CHNS cohort with access to health care and the existence of withdrawal bias. Besides, different criteria for recognizing overweight and obesity in children may influence the accuracy of the estimation.

## 5. Conclusions and Implications

Our study identified that several adiposity indices—namely BMI, WC, HC, TST, and BRI— were correlated with a heightened risk of hypertension among Chinese children and adolescents. Notably, only BMI demonstrated a consistent, directional relationship with the longitudinal presentation of hypertension symptoms. Moreover, the pronounced associations between these adiposity indices and the risk of hypertension were predominantly observed in the younger age group, ranging from 7 to 12 years old.

## Acknowledgements

The authors thank the CHNS team for their hard work and unselfsh sharing of survey data. This research uses data from China Health and Nutrition Survey (CHNS). We are grateful to research grant funding from the National Institute for Health (NIH), the Eunice Kennedy Shriver National Institute of Child Health and Human Development (NICHD) for R01 HD30880, National Institute on Aging (NIA) for R01 AG065357, National Institute of Diabetes and Digestive and Kidney Diseases (NIDDK) for R01DK104371 and R01HL108427, the NIH Fogarty grant D43TW009077 since 1989, and the China-Japan Friendship Hospital, Ministry of Health for support for CHNS 2009, Chinese National Human Genome Center at Shanghai since 2009, and Beijing Municipal Center for Disease Prevention and Control since 2011.

## Abbreviation

AUC: Area Under Curve;
BMI: Body Mass Index;
BP: Blood Pressure;
BRI: Body Roundness Index;
BSI: Body Shape Index;
CHNS: China Health and Nutrition Survey;
CI: Conicity Inex;
CMRFs: Clustered Cardio-metabolic Risk Factors;
CVD: Cardiovascular Disease;
eGFR: estimated glomerular filtration rate;
HC: Hip Circumference;
HDL-C: High-density Lipoprotein Cholesterol;
IDI: Integrated Discrimination Improvement;
IQR: Interquartile Range;
NRI: Net Reclassification Improvement;
SD: Standard Deviation;
SUA: Serum Uric Acid;
TG: Triglycerides;
TST: Triceps Skinfold Thickness;
VFI: Visceral Fat Index;
WC: Waist Circumference;
WHR: Waist to Hip Circumference Ratio.

## Author contribution

BZ, SZ, and JF contributed to the conception or design of the work. BZ, HS and XW contributed to the acquisition, analysis, or interpretation of data for the work. BZ and RW drafted the manuscript. JY and XG critically revised the manuscript. BZ and TW contribute to the analysis or interpretation of the work. All gave final approval and agreed to be accountable for all aspects of work ensuring integrity and accuracy.

## Ethical approval

The China Health and Nutrition Survey (CHNS) was granted approval by the Institutional Review Board at the University of North Carolina at Chapel Hill, as well as by the local Institutional Review Board of Ethics Committee. As this study is based on secondary data analysis of an existing dataset, we did not require additional ethical approval from our institution. Original data generated and analyzed during this study are included in this published article or in the data repositories listed in References.

## Data Availability

Original data generated and analyzed during this study are included in this published article or in the data repositories listed in References.

